# When Care Depends on the Caregiver: Lived Experiences of Latino Families Navigating Dementia Care Pathways

**DOI:** 10.64898/2026.03.29.26349413

**Authors:** Maria Mora Pinzon, Rosie Pasqualini, Veronica Navarro, Maria del Carmen Rosales, Olivia Franzese, Jaime Perales-Puchalt

## Abstract

**Introduction:** Latino families shoulder a disproportionate share of dementia care in the United States, yet encounter multilayered barriers that shape access, timeliness, and quality. This study explores the experiences of Latino care partners, focusing on how system-level, cultural, and linguistic factors shape dementia care.

**Methods:** We conducted a qualitative study using semi-structured interviews with care partners of Latino individuals living with Alzheimer’s disease and related dementias (ADRD). Interviews were conducted by phone or videoconference by a bilingual interviewer, and the interviews were recorded and transcribed verbatim. Data was analyzed using reflexive thematic analysis.

**Results:** Twenty-three participants were recruited. Two meta-themes captured participants’ experiences. (1) Mismatch Between the Healthcare System and the Lived Realities of Latino Families Affected by Dementia, which included three subthemes: a) Linguistic barriers that referred to the quality and dialect fit (over-literal jargon, unfamiliar regional vocabulary, poor adaptation to literacy); b) Cultural misfit, were dementia-care programs were not culturally or linguistically appropriate, or programs where cultural norms were disregarded; and c) Structural and systemic barriers, such as communication failures (e.g. voicemail loops, no responsiveness) and long waits/fragmented pathways that broke clinical momentum (e.g. months to a year for specialty appointment). The second theme was: The Central Role of the Latino Caregiver in Navigating Dementia Care, where, in the absence of pathway ownership, care partners served as navigators, interpreters, coordinators, and safety monitors, while also bearing the emotional and financial strain.

**Discussion:** The narratives from care partners reveal specific mechanisms (e.g., caregiver hyper-advocacy and “maze-like” coordination failures) that, if addressed, can guide intervention design and policy aimed at redistributing coordination back to the system and improving outcomes for Latino families.

## INTRODUCTION

Caregiving refers to providing regular care or assistance to relatives or friends living with chronic illness or disability. As of 2025, an estimated 63 million U.S. adults serve as family caregivers for an adult or child with a medical condition or disability, showing a sharp rise since 2015.^1^ Within Latino communities, the most recent data estimates indicate that Hispanic/Latino caregivers comprised approximately 17% of U.S. adult care partners in 2020,^1^ which translates to 10 million Latino caregivers nationally.

Alzheimer’s disease and related dementias (ADRD) unfold within social, cultural, and structural contexts, meaning that the health outcomes of patients and their care partners cannot be separated from the environments in which they live. Community resources, policy structures, and social attitudes shape access, diagnosis, and management, and these influences are especially pronounced in Latino communities, where cultural norms (e.g., familism and personalism), socioeconomic constraints, and structural inequities intersect to intensify both risk and caregiving burden.

Because family and informal caregivers play multifaceted roles, they require a suite of supports (education, navigation, respite, transportation, and culturally/linguistically tailored services) to remain healthy and sustain care.^2,3^ Yet even when programs exist, they are frequently under-utilized due to implementation barriers, complexity, transportation and scheduling obstacles, and limited awareness.^3–6^ But few studies trace the full dementia care pathway from Latino families’ perspectives, and limited qualitative work centers the lived experience across settings and time. Addressing these gaps requires proactive, culturally responsive outreach and navigation-first models that operationalize language/dialect fit and family partnership.

This qualitative study explores the experiences of Latino families navigating dementia care, focusing on how system-level, cultural, linguistic, and relational factors shape access, diagnosis, and caregiving. We present findings on the misalignment between families’ lived experiences and the healthcare system, the central, often overwhelming responsibilities assumed by care partners. These results can inform the design and implementation of dementia care models that are accessible, timely, and culturally/linguistically congruent for Latino families.

## METHODS

### Design, Population & Recruitment

This qualitative study planned to identify what ADRD care services are offered in primary care and how they are delivered across a variety of settings. Care partners were recruited via convenience sampling from diverse sources, including a research registry, clinics, community ADRD patient lists, internet and newspaper advertisement. The research team also leveraged connections with community partners and team members built since 2015.

### Data Collection & analysis

After participants provided informed consent, interviews were conducted by a bilingual researcher via telephone or a secure videoconference platform. All interviews were audio-recorded with permission, professionally transcribed verbatim, and checked for accuracy. Transcripts were then de-identified for data analysis. We used a reflexive thematic analysis approach.^7,8^ Four reviewers independently coded an initial subset of transcripts to develop a shared codebook, meeting regularly to refine code definitions. The full dataset was then coded using the codebook, and themes were further refined through team discussion and organized into higher-order categories (meta-themes), with representative quotes selected to illustrate each theme.

### Ethics/Institutional Review Board Review

The data was collected at the University of Kansas Medical Center and analyzed at the University of Wisconsin-Madison. The University of Kansas Medical Center Institutional Review Board approved the project (STUDY00145615). Participants provided informed written consent online. The University of Wisconsin-Madison Health Sciences Institutional Review Board determined that the research met exemption criteria. A Data User Agreement facilitated the sharing of de-identified data between institutions.

## RESULTS

Of the 21 participating care partners, 19 were women (90%), 16 were younger than 66 years (76%), and 15 were adult children (71%) of the care recipients, 14 of whom had Alzheimer disease (67%). All but one participant identified as Latino with 6 (29%) being born in the United States; and all lived in urban areas, mostly in the Midwest (n=15; 71%). Interviews were conducted in Spanish with 13 (62%) caregivers, 14 (67%) of them. Figure 1 shows our two overarching themes and the subthemes identified, along with a sample quote for each. For each subtheme, we provide participant quotations followed by the corresponding participant number.

### Theme 1. Mismatch Between the Healthcare System and the Lived Realities of Latino Families Affected by Dementia

Across interviews, Latino care partners described a persistent mismatch between what they needed to care for a loved one with dementia and what health and social service systems provided. Care partners consistently linked this mismatch to linguistic, cultural, and structural gaps that complicate accurate assessment, delay care, and place additional burdens on families.

### 1.1 Linguistic and Interpretation Barriers

Families repeatedly emphasized that high-quality interpretation services that accounted for patients’ dialects and characteristics were necessary for accurate assessments and adequate care. Two interlocking subthemes emerged in this category:

#### Interpreter quality problems

Care partners described interpreters who translated literally without ensuring understanding, used overly specialized vocabulary, or were inconsistently available. Importantly, several care partners perceived that some interpreters were unable to adapt explanations for a patient with limited formal education. At these moments, care partners stepped in to “fix” the interaction, which they felt kept the visit moving but also shifted the burden of clinical communication onto the family. As one caregiver explained:

> “The people who translate don’t translate in a way that you can understand; they use very complicated words” (CP 1 - Translated from Spanish)

Another caregiver described how the interpreter’s limitations collided with the patient’s educational background:

> “My mother’s educational level is very low, so many of the interpreters didn’t understand her; I had to intervene and explain things to them” (CP 3 - Translated from Spanish)

### Dialect mismatch

Beyond general interpreter quality, families highlighted dialect as a critical but often overlooked dimension of language access. Even when Spanish interpretation is provided, regional variation in vocabulary and idioms can impede mutual understanding, especially for patients with rural backgrounds or limited literacy, or those who may rely on familiar phrasing to achieve understanding. Care partners linked dialect mismatches to confusion, distress, potential for misdiagnosis, and even accelerated decline. One caregiver noted:

> “Now there’s one thing about translations, many times they’re not done well. Or they’re with [different] words; here the population is often Mexican, so you know that sometimes some things vary a little, I mean, the meaning of some things” (CP 2 - Translated from Spanish)

Families articulated the need for dialect-specific matching, and on occasions, how they would like to intervene to ensure adequate understanding.

> “There is a different dialect for every country, I mean we all speak Spanish, but things mean different things to people from different parts of Latin American countries.” (CP 16)

> “They have never allowed any of the family members to be there when she is doing her testing. The first couple of times, she just did it on her own without an interpreter, which I asked you know, she would understand better to know what to do if we helped her and talked to her, but they did not want us to help her.” (CP 16)

### 1.2 Cultural Misfit

Care partners consistently described how dementia services do not fit with Latino cultural and language contexts. This misfit was shown on two sub-themes: programs were not culturally or linguistically congruent, or cultural norms were poorly understood or disregarded. Together, these gaps shaped how families found (or failed to find) services, whether elders felt respected and engaged, and whether families trusted formal systems enough to continue seeking support.

#### Programs not culturally/linguistically congruent

Care partners reported that locating dementia-specific resources in Spanish was time-consuming, difficult in certain areas, and often unsuccessful. Even when services existed, they were not necessarily dementia-informed or designed around cognitive impairment, resulting in activities that elders could not follow or enjoy. One caregiver described the scarcity as follows:

> “It took me a very long time to find resources that were available for Spanish speakers, and it is only in certain communities… you know, senior centers, they are just completely lack of any resources in Spanish, and that was one of the most difficult things that I have found through this journey” (CP 15)

Other services were inconsistent with cultural expectations around food and daily routines, diminishing acceptability. As one participant explained when asked about a home-delivered meal program:

> “We had meals on wheels, but we just could not eat the meals; they were terrible. They were terrible for a [LATIN AMERICAN] family” (CP 12)

Another care partner described the downstream impact when no one could understand the patient in Spanish at the long-term care facility:

> “No one understood what he was saying [whether it was at a hospital or residential facility]. It was literally heartbreaking, and I would say that it was why he was so quickly declining… that he could not be understood” (CP 15)

#### Unrecognized Cultural Norms

Care partners also described routine encounters in which cultural values were misinterpreted or dismissed. A recurring example was the display of disrespect by staff, who talked down to older adults or infantilized them. One care partner recalled their parent’s reaction in a clinical encounter:

> “Many times I heard my dad say frustrated, ‘Why does she have to talk to me like a child? I understand perfectly well what she’s saying.’” (CP 2, translated from Spanish).

Care partners emphasized the preference for home-based care, sometimes with informal helpers, and strong hesitancy toward institutionalization. When this norm was not understood, conflict with providers and social services followed:

> “In our countries, we try to have helpers, even if they aren’t qualified, to have them by the elderly person side always, all the time. So, they don’t understand that, and I had several arguments with the social worker.” (CP 5 - Translated from Spanish)

Experiences inside facilities reinforced these concerns when staff relied on token Spanish or an infantilizing tone:

> “The nursing home [staff] were mostly all English speaking… and you would hear the “mamita”, and [they] come out with stupid little Spanish words. That lady [Patient], she’s old, she is confused, can you give her a sentence? Because you would hear them, they would tell her no!, no!, no! like she was a child, like don’t do this or don’t do that, and I said, if I was a secret shopper, all these people would be fired because they were not doing their job to the best of their ability” (CP 19)

### 1.2 Structural and Systemic Access Barriers

Care partners described routine, system-level obstacles that made even basic tasks, such as getting a refill, feel unpredictable and exhausting. Two interlocking subthemes emerged: system-wide communication failures and long wait times with fragmented pathways. Together, they generated a pervasive sense that the healthcare system was hard to reach, slow to respond, and poorly coordinated, leaving families to “chase” the system.

#### System-wide communication failures

Care partners repeatedly encountered voicemail loops, unreturned calls, and nonfunctioning “direct” lines, requiring them to escalate their attempts to communicate (e.g., messages, phone calls, portal notes, contacting assistants) just to obtain a response. One caregiver described the following experience:

> “We left that day [after the appointment], and I was waiting for the order. That was a Thursday, by Tuesday I had no answer as to whether they had sent an order. I wrote to her, I called, I wrote to her, I spoke with her assistant, with the doctor, and then they answered me the next day [Wednesday]” (CP 1 - Translated from Spanish)

Even numbers offered as “direct support” failed in practice:

> “Getting to speak to a live person [in neurology] is almost [sigh]. The other day I had a Zoom with them, and the girl said I am going to give you my direct number in case you can’t figure out what to do with Zoom. I did have a problem, I called that number she gave not even five minutes earlier and no answer.” (CP 16)

These events were common enough that care partners describe these experiences as anticipated rather than one-of instances:

> “You cannot talk to anyone, you call the main number and they say, “this is so and so, we will get back to you in one or two days”. Well, it is seven days and no message, we leave the same message again and then again, and nobody returns phone calls.” (CP 16)

#### Long wait times and fragmented pathways

care partners faced waits of weeks to a year for specialty visits. Care was described as “scattershot”, with little input or participation of primary care providers, and no proactive follow-up. As explained by one care partner, navigating a large health system was a barrier in itself:

> “Probably the biggest thing is navigating through [CLINIC], and that is a challenge in and out of itself.” (CP 17)

Several care partners noted the diminished role of primary care in dementia care, with some describing limited guidance or linkage to resources:

> “The primary care doctor, all she does is refer. She doesn’t know what programs are available, she doesn’t say, ‘Go to this place,’ she doesn’t say, ‘Wait for this.” (CP 7 - Translated from Spanish)

> “She has been a patient for the last three to four years, and I am surprised they still have not called. So we found some other resources, and I do not understand how a person in their 80’s has not gotten a call from her primary place.“ (CP 16)

Long waits, particularly to get an appointment with Neurology, were a frequent complaint, with tangible consequences, such as missed care:

> “In neurology, appointments are very difficult to get. They’re very spaced out, and many times you have to wait up to a year for an appointment. If you miss one, then you lose the sequence.” (CP 2 - Translated from Spanish)

### Meta Theme 2. The Central Role of the Latino Caregiver in Navigating Dementia Care

Across interviews, care partners described taking on multiple system-facing roles because healthcare, social services, and community systems lacked the coordination, language access, or proactive services needed to support dementia care. Rather than simply providing emotional and daily support, care partners became the primary navigators, interpreters, advocates, care coordinators, and financial buffers for their loved ones. The result was an intensely demanding form of caregiving that changed family dynamics and often determined whether or not a loved one received timely and appropriate care.

### 2.1 Care partners as Navigator, Interpreter, and Advocate

Families described a constant need to push the system forward. These efforts were described as exhausting but necessary to prevent care stagnation or miscommunication. Within this theme, three related roles were identified as needed to execute the caregiver role: advocate, interpreter, and care coordinator.

#### Hyper-advocacy

Care partners repeatedly emphasized that, without persistent pressure, care simply did not happen. As a result, care partners felt responsible for “making the system listen.” One caregiver expressed the strain of repeatedly clarifying and restating concerns:

> “Because I feel like I have to repeat myself all the time… I feel like that’s where I get the most gray hairs, because it’s like we’re speaking two different languages” (CP 11 - Translated from Spanish)

Others described having to explicitly argue for specialist evaluation because primary care was not recognizing the urgency:

> “I told her, I need to see a specialist who can tell me what’s going on, because I see all these things, and I don’t know what she has. I can’t keep treating her like she just has the flu or something, I told her. There’s something more.” (CP 5 - Translated from Spanish)

Advocacy also extended to monitoring the timeline of care:

> “Every time I took her to the neurologist, I had to ask… Today I sent a message to the coordinator, to the director, and I said, I have to talk to the doctor or the nurse practitioner and tell them it’s time to go to the doctor again, because it’s been two years since we had an evaluation.” (CP 5 - Translated from Spanish)

#### Acting as de facto interpreter

Care partners also stepped into the interpreter role due to absence, inconsistency, or inadequate professional interpretation. This responsibility was on occasion described as unavoidable:

> “Two or three years ago, there were hardly any translators here, so we translated ourselves” (CP 7 - Translated from Spanish)

> “[A translator] was not offered, I think just because they knew I was there, it was not offered.” (CP 15)

#### Care coordination

Care partners described the time and effort that they invested in connecting their loved ones to community resources, specialty care, insurance benefits, and safe daily routines. With no centralized pathway, these responsibilities required repeated calls, extensive searching, and trial-and-error navigation through unfamiliar systems. One caregiver detailed the long process of locating appropriate dementia-specific supports:

> “I struggled so much trying to find places that were specific care for Alzheimer’s patients, not just a regular senior center. I do not know how many phone calls I had to make, and it took me two years to finally find a place, if not more, maybe three years, to find a place for my mother to go to a senior center that was specific for Alzheimer’s patients.” (CP 15)

Others avoided potentially helpful programs because they lacked trusted guidance:

> “I have never really explored [services like Meals on Wheels], I always felt they were sort of income-related, and I was always afraid that we would not qualify for anything, so I just never explored that sort of help really.” (CP 14)

Furthermore, the “support” given was often generic and mismatched to their needs, and care partners had to further navigate these services alone, making the process feel unplanned and unsupported:

> “The social worker [said] here is a list of you know available services, and those services are usually the how to understand Alzheimer’s, what to watch out for; when I felt that the needs that we had were not being addressed by those. I think the information is great, but I think families have a real, real need for things like helping make appointments, looking for respite care programs, making those phone calls, looking for residences, looking for programs.” (CP 15)

### 2.2 System-Induced Caregiver Financial Burden

Care partners frequently shouldered significant financial strain because publicly available support was insufficient, inconsistent, or poorly matched to their needs. Many families attempted to supplement care to maintain safety and dignity at home, yet the system offered limited practical support for doing so. One caregiver shared:

> “…so I had to quit my job to be there for him, to take care of him.” (CP 13, Translated from Spanish)

Another described continual financial pressure:

> “You look for the services the government provides, but they aren’t enough for everything we need at home, so you’re constantly digging into your own pocket” (CP 7 - Translated from Spanish)

## DISCUSSION

Our results show that Latino families face multilayered, mutually reinforcing barriers to securing dementia care. At the patient-caregiver interface, literal or jargon-heavy interpretation and dialect mismatches impede comprehension. At the same time, the absence of clear navigation roles pushes caregivers to act as de facto interpreters, navigators, and coordinators. Within care settings, chronic communication breakdowns, long waits, and fragmented pathways force families to “chase” care, risking missed or discontinued services. In the community, scarce dementia-specific Spanish-language programs and services misaligned with cultural foods, activities, or literacy limit engagement and add financial strain as care partners fill gaps. Taken together, these barriers determine whether care is accessible, timely, and culturally/linguistically congruent, ultimately making caregiver effort-- not system design-- the decisive factor in care quality.

Although language and interpretation barriers for Latino patients are well documented,^3,4,9,10^ this work adds to prior research by describing how care partners evaluate not just the presence but the quality of interpretation, especially the interpreter’s familiarity with the patient’s specific dialect. The care partners in this study show that interpreter-mediated encounters can fail to achieve true comprehension when interpreters use specialized jargon, translate verbatim without ensuring understanding, or are unfamiliar with regional dialects. These results are supported by recent qualitative studies with Spanish-speaking patients and caregivers, which document medical jargon and dialect mismatches as central drivers of miscommunication and the perceived need for families to “re-interpret.”^3,4,11,12^ We also identify a concern from care partners, which linked interpreter quality to threats to diagnostic validity in cognitive testing, resulting in caregivers’ desire to step in to “fix” instructions, a practice that is not described within neuropsychological assessment, and which prompt concerns for accurate performance on the tests.^13^

Our findings also reflected the lack of culturally appropriate services at the community level. Community-based programs can improve outcomes for people with dementia and their care partners. However, programs that are not linguistically accessible, dementia-specific, or culturally aligned are less likely to be adopted, leading families to disengage or assume full responsibility for care.^14–16^ This was also shown by Martinez et al. In their study, they described the “culture trap” as the assumption that individuals will have a broad family safety net and be interested in caring for individuals at home, which results in under-referral to existing services.^3^ However, it is important to consider that evidence from community-engaged initiatives shows that simple availability is not enough; programs need to be culturally tailored and have proactive navigation/coordination for sustained engagement.^3,17^ Yet a persistent design gap remains: interventions for Hispanic caregivers rarely incorporate intersectional factors such as socioeconomic constraints, literacy levels, sexual and gender minority identities, or disability.^18^ For programs to be effective and equitable, they must integrate language/dialect fit, caregiver navigation support, and intersectional tailoring from the outset, rather than adding cultural elements superficially.^19^

An important aspect of our study is the care partners’ thoughts on communicating with older adults, in which using infantilizing “elderspeak” language and broken Spanish were considered disrespectful and contrary to the cultural value of *“respect”.* To our knowledge, there is limited discussion of this topic among Hispanic/Latino older adults in the literature. These findings correspond with the existing literature, which increasingly recognizes this speech as disrespectful because it undermines personhood. Observational work in U.S. nursing homes shows that elderspeak, which is described as baby-talk features such as diminutives (“good girl”), collective pronouns (“are we ready for our bath?”), exaggerated pitch, and simplified grammar are perceived by older adults as patronizing and demeaning rather than supportive, while also increasing resistance to care among residents with dementia.^20–22^

Our data portrays a maze-like communication environment that stalls clinical progress and shifts administrative work onto families, including voicemail loops, unreturned portal messages, and delayed orders, among others, which breaks clinical momentum when continuity matters most. Similar patterns have been described in other studies, with fragmented transitions and long waits disrupting the pathway to diagnosis and follow-up.^23,24^ Evidence from provider surveys also maps where momentum breaks: PCPs’ confidence in dementia workflows is modest, and although referral processes are variable, this results in inconsistent action on screening results.^25,26^ In line with national caregiving data, more families now struggle to coordinate care than five years ago, underscoring how disjointed systems shift coordination onto caregivers.^1,27^ Our findings contribute to the literature by showing how communication failures and long waits mutually reinforce each other for Latino families already navigating linguistic and cultural misalignment, amplifying missed assessments, medication lapses, and role reconfiguration in which routine system tasks are outsourced to families.^28^ Collectively, the evidence suggests that improving “access” requires reliable, reachable communication infrastructure (including after-hours bilingual capacity), proactive pathway ownership (a single key contact or coordinator), so that clinical momentum is not lost to the system’s own friction.

Our findings further show the consequences of fragmented, hard-to-reach dementia care: care partners become the central organizing force, taking on the role of connecting siloed organizations, securing resources, managing logistics, and problem-solving across agencies, which we called the “hyper-advocacy burden.” Qualitative work with Latino caregivers reinforces this, a need to coordinate care across settings,^29^ while also navigating cultural incongruence, leading families to avoid formal services and shoulder more tasks at home.^3,15,28^

Most of the dementia-care literature foregrounds the psychological burden on family caregivers’ stress, depression, and emotional strain; which our participants also described, even if we did not present those data here (e.g., feelings of exhaustion, grief, and isolation).^16,30–33^ By contrast, coordination-related burdens are typically examined from the healthcare side, for example, surveys and implementation studies that document limited clinician time, variable referral practices, and workflow/follow-up gaps.^9,25,34^ Our results add to this literature by linking the two: when systems lack pathway ownership and reachable communication, care partners are forced into expanded roles, on top of psychological load.

A limitation of this qualitative study is that it is based on a small number of interviews, which allows for depth but limits breadth; the findings reflect the experiences and contexts of those who participated and should not be generalized to all Latino families living with dementia in the U.S. As with most volunteer samples, there is a risk of selection bias, where care partners who consented to being interviewed may differ systematically from those who did not. These accounts may underrepresent the most marginalized families, such as those not connected to services, those with limited phone or internet access, those facing immigration-related concerns, or those who avoid research due to mistrust or fear. Even with these constraints, the narratives consistently converged on a series of themes that open new lines of research and future intervention development.

### Implications for Practice and Policy

For Latino and Hispanic populations in particular, effective interventions must operate across multiple levels: strengthening caregiver supports and culturally grounded services at the community level, ensuring accessible policies at the organization level, and addressing stigma and socioeconomic disparities at the societal level. No single intervention, whether clinical, educational, or policy-based, can adequately address the multiple challenges faced by patients and caregivers. Instead, improving outcomes requires coordinated efforts that recognize Alzheimer’s disease as embedded within social, cultural, and economic systems.

Some specific solutions implied by the results of our research include: ensuring that communication systems are reachable, reliable, and dialect-attuned (including after-hours bilingual access), using a single named navigator to manage and coordinate referrals and follow-up, and funding interpreter training tailored to regional dialects and literacy. Furthermore, enhancing the availability of bilingual, dementia-specific community supports (not generic “Spanish services”). These changes shift care from relying on caregiver hyper-advocacy to proactively delivered, equitable support.

The literature also points to several important directions for future research. Longitudinal studies are needed to understand how culturally specific caregiving dynamics evolve, particularly in Latino families, where intergenerational expectations and acculturation that shape caregiving roles might change over time. Community-based interventions should be evaluated across diverse populations to determine whether their benefits generalize or require cultural tailoring.

## Conclusion

Our findings and the literature argue for an integrated redesign: culturally and linguistically aligned cognitive assessments, interpreter training in regional dialects and literacy-adapted communication, embedded navigation roles to assume coordination work the system currently offloads to families, reliable phone/portal infrastructure with reachable bilingual support, and bilingual dementia-specific community programs that families will actually use. Implementing these elements and measuring success by reach, timeliness, continuity, and caregiver workload will shift dementia care from a system that depends on caregiver hyper-advocacy to one that proactively delivers equitable, trustworthy, and sustainable support for Latino families.

## Data Availability

Data is available upon reasonable request to the corresponding author at.

---

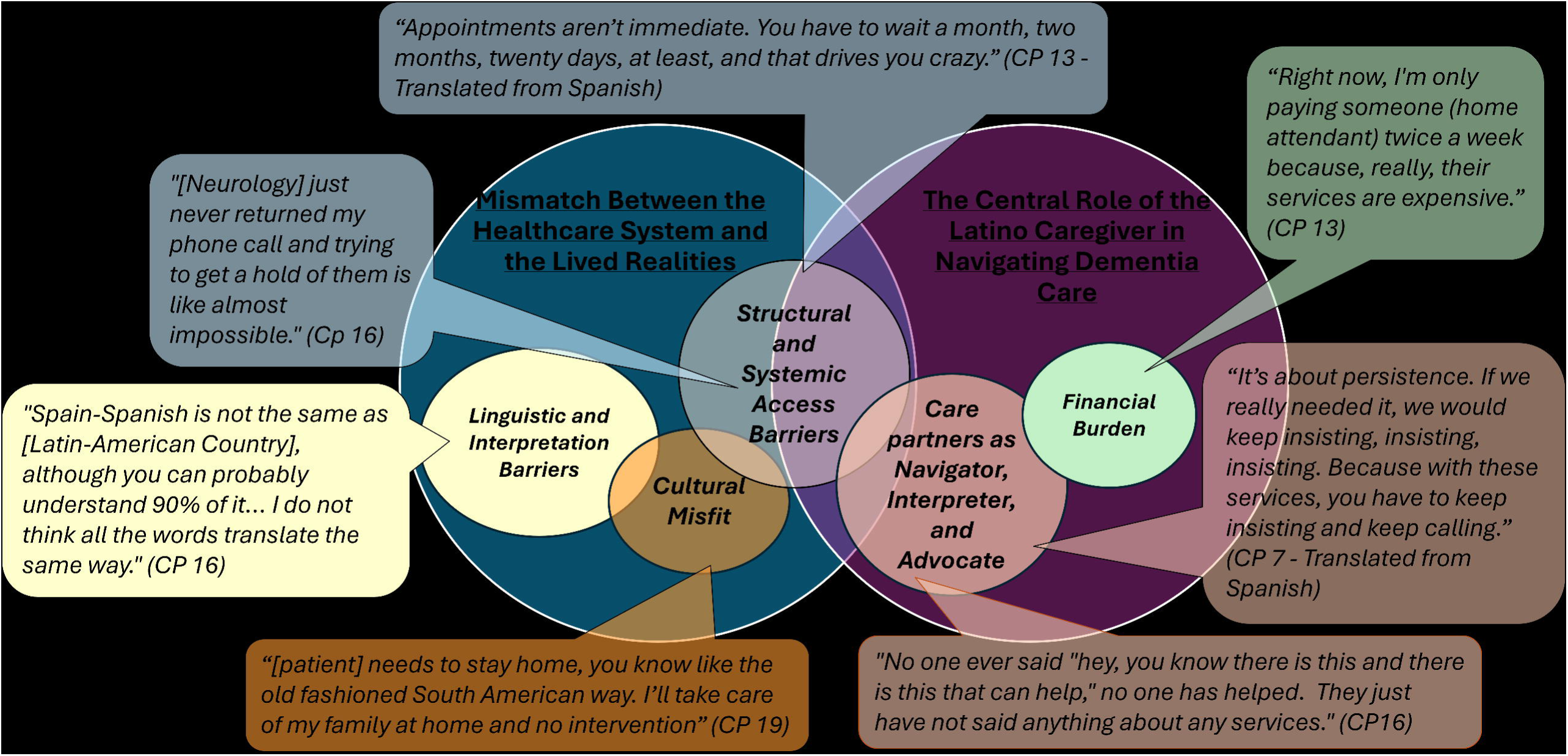

